# Distinct Resting-State Functional Connectivity Profiles in ADHD with and without Prenatal Alcohol Exposure

**DOI:** 10.64898/2026.05.25.26354061

**Authors:** Ishika Gupta, Lea Farkouh, Lisa Kilpatrick, Jennifer Korthas, Noriko Salamon, Benjamin N. Schneider, Shantanu H. Joshi, Jeffry R. Alger, Mary J. O’Connor, Joseph O’Neill

## Abstract

**Aim:** To determine whether the neural phenotype (whole-brain resting-state functional connectivity pattern) of attention-deficit/hyperactivity disorder associated with prenatal alcohol exposure (ADHD-PAE) differs from that in unexposed children with ADHD of probable familial origin (ADHD-PAE).

**Method:** Resting-state functional MRI was acquired from 26 children with ADHD+PAE, 25 with ADHD-PAE and 25 typically developing (TD) children, all aged 8-13 years. Mean connectivity matrices based on the Cole-Anticevic Brainwide Network Parcellation of the brain were compared between the groups.

**Results:** Within the frontoparietal network (FPN), children with ADHD+PAE showed widespread lower group-mean connectivity than children with ADHD–PAE; effects were concentrated primarily in cerebellar–cerebral cortical and cerebral cortical–cerebral cortical connections. Children with ADHD–PAE showed widespread hyperconnectivity relative to TD children. Children with ADHD+PAE showed mixed hyper- and hypoconnectivity relative to TD.

**Interpretation:** These results are consistent with other MRI findings indicating that ADHD+PAE is neurally distinct from ADHD-PAE; PAE may be associated with broadly reduced connectivity, especially across cerebellar–cerebral cortical systems.

## 1. Introduction

Attention deficit hyperactivity disorder (ADHD), the most common neurodevelopmental disorder in childhood (Yang et al. 2022), arises from diverse etiologies (Kian et al. 2022). One major etiology is prenatal alcohol exposure (PAE), which substantially increases the risk of ADHD symptoms and related cognitive and behavioral difficulties (Fryer et al. 2007, O’Connor 2014, San Martin Porter et al. 2019). In clinical practice, children whose ADHD is associated with PAE (“ADHD+PAE”) are often misdiagnosed as unexposed children whose ADHD is due to other, most often familial, causes (“ADHD-PAE”; Young et al. 2016, Mattson et al. 2013, O’Connor 2014), whereby the underlying PAE goes unrecognized. Such misdiagnosis has clinical consequences. Psychostimulants are used as first-line therapy for both ADHD-PAE and ADHD+PAE. But in ADHD+PAE, these drugs may be less effective (Doig et al. 2008, Infante et al. 2011, Oesterheld et al. 1998, Snyder et al. 1997, Young et al. 2016) and may induce side effects at higher rates (Coe et al. 2001, Ipsiroglu et al. 2015, Ji & Findling 2016). Long-term prognosis also differs between these two ADHD etiologies. As children age into adolescence and adulthood, symptoms gradually resolve with or without treatment for many with ADHD-PAE (Jensen et al. 2007), but people with ADHD+PAE more typically continue to evince problems in attention and executive function, behavioral and emotional regulation, and adaptive function (O’Connor 2014, Streissguth et al. 1996, Young et al. 2016). These “secondary disabilities” of ADHD+PAE contribute to adverse long-term outcomes (O’Connor & Paley 2009, Streissguth et al. 1996), including school and legal trouble, substance abuse, mental illness, and suicide (Burd et al. 2007, Fast & Conry 2004, O’Connor 2014, O’Connor et al. 2019, O’Connor & Paley 2009). Thus, presence of PAE is highly relevant to the management of ADHD.

The above differences in treatment response and prognosis suggest that partially distinct brain mechanisms underlie ADHD+PAE and ADHD-PAE (Alger et al. 2021, O’Neill et al. 2019). Our past work with cortical gyrification, intracortical myelin mapping, diffusion tensor imaging (DTI), and proton magnetic resonance spectroscopy (MRS) supports the notion that the pediatric brain notably differs in ADHD+PAE versus ADHD-PAE and that such differences may be used to classify individual patients by etiology (Alger et al. 2021; Kilpatrick et al. 2021,2022; O’Neill et al. 2019, 2022). This raises a key question: does ADHD+PAE reflect the same neural phenotype as ADHD-PAE, or is it a neurally distinct subtype with different circuit-level alterations?

Neuroimaging work in fetal alcohol spectrum disorders (FASD) has begun to address this question by characterizing large-scale brain networks. Resting-state fMRI (rsfMRI) studies have shown that children with FASD exhibit global functional connectivity abnormalities and reduced network efficiency, which relate to cognitive deficits such as attention and executive functioning problems (Joshi et al. 2013, Wozniak et al. 2017). Using independent component and network-based approaches, it was found that children and adolescents with PAE show both hypo- and hyperconnectivity across multiple resting-state networks compared with controls; these connectivity differences were associated with motor, learning, and memory impairments (Little et al. 2018). In neonates, Donald et al. (2016) found altered interhemispheric functional connectivity and atypical coupling between the somatosensory, motor, brainstem, and thalamic networks which indicates that intrinsic network organization is disrupted very early following PAE. More focal analyses have shown that there are localized reductions in resting-state connectivity within cognition-related networks in children with PAE, with lower connectivity partially mediating poorer arithmetic performance (Fan et al. 2017,2024). Task-based fMRI studies also report altered fronto-parietal connectivity during spatial working memory and atypical recruitment of default mode and attention networks during cognitive tasks in youth and adults with PAE (Infante et al. 2017; Santhanam et al. 2009, 2011). Yet, despite this growing literature, no study has directly compared whole-brain resting-state functional connectivity in children with ADHD+PAE, ADHD-PAE, and typically developing (TD) controls.

In the present study, we address this gap using high-quality, Human Connectome Project–style rsfMRI (Smith et al. 2013) and the Cole-Anticevic Brainwide Network Parcellation (CAB-NP) pediatric brain atlas (Ji et al. 2019) in a well-characterized sample of 8–13-year-olds. We scanned three groups: children with ADHD and documented prenatal alcohol exposure (ADHD+PAE), children with ADHD and no evidence of PAE (ADHD–PAE, presumed primarily familial etiology), and typically developing controls (TD). Our core question was whether children with ADHD+PAE are neurally distinct from ADHD-PAE and TD children. We investigated whether ADHD+PAE shows a distinct whole-brain connectivity signature, instead of just representing an amplified version of the ADHD-PAE pattern. We hypothesized that ADHD–PAE would show a predominantly hyperconnected profile relative to TD, whereas ADHD+PAE would be characterized by a more globally underconnected architecture, consistent with prior PAE and FASD work.

## 2. Methods

### Participants

Participants were recruited and assessed as in our prior work (Alger et al. 2021; Kilpatrick et al. 2021, 2022; O’Neill et al. 2021). Briefly, recruitment sources included FASD organizations, national websites, other pediatric studies, and physician referrals. Candidate participants were screened by telephone. Eligible participants underwent demographic and medical history, diagnostic and clinical interviews, and mock scanning, then actual MRI scanning. Children taking stimulants were asked to be off medication for at least 24 hr prior to testing and scanning. The UCLA Institutional Review Board approved all procedures. All parents gave informed consent; all children gave assent and were compensated.

Seventy-six children between 8 and 13 years of age with full-scale IQ (FSIQ) ≥ 70 (WASI-II; Wechsler 2011) participated in the study (Table 1). The participants were divided into three groups: children with attention deficit hyperactivity disorder and confirmed prenatal alcohol exposure (ADHD+PAE; n = 26), children with ADHD and no evidence of prenatal alcohol exposure (ADHD–PAE; n = 25), and typically developing controls (TD; n = 25). These were the final numbers analyzed after exclusions and quality assurance of MRI data (see below). Children in the ADHD+PAE group had to meet criteria for fetal alcohol syndrome (FAS), partial fetal alcohol syndrome (pFAS), or alcohol-related neurodevelopmental disorder (ARND) using the modified Institute of Medicine (IOM) criteria (Hoyme et al. 2016; see O’Connor et al. 2019). Prenatal exposure to alcohol and other teratogens was assessed using the Health Interview for Women (HIW; O’Connor & Kasari 2000) or the Health Interview for Adoptive and Foster Parents (HIAFP; Quattlebaum & O’Connor 2013). Criteria for alcohol exposure included >6 drinks/week for ≥2 week and/or ≥3 drinks on ≥2 occasions including prior to and following pregnancy recognition. For adoptive/fostered participants, data regarding prenatal exposure to alcohol and other teratogens were obtained via birth, medical, or adoption records, or by reliable informants. This approach is considered acceptable for establishing PAE by the scientific community (CDC, 2004). Children in the ADHD-PAE group had to have one or more first-degree relatives with diagnosed ADHD to support a probable genetic etiology of ADHD. Children in the ADHD groups met DSM-5 criteria for ADHD, any subtype, according to the Schedule for Affective Disorders and Schizophrenia for School-Aged Children Parent Version (K-SADS P; Kaufman et al. 1997, Townsend et al. 2020). Children in the ADHD-PAE group had to have one or more first-degree relatives with diagnosed ADHD to support a probable genetic etiology of ADHD and exposure to <2 standard drinks (1.20 oz absolute alcohol) throughout gestation. Children in the TD group were required to have no first-degree relative with diagnosed ADHD and exposure to <2 standard drinks throughout gestation. Severity of ADHD was assessed by Parent ratings on the Conners 3 Behavior Rating Scale (Conners 2008).

**Table 1.** Final sample demographic and clinical variables.

|  | ADHD+PAE | ADHD-PAE | TD |
| --- | --- | --- | --- |
| **N** | 26 | 25 | 25 |
| **Sex, male/female** | 14/12 | 18/7 | 12/13 |
| **Age, years** | 9.7±1.3 | 10.4 ±1.3 | 10.4 ±1.4 |
| **Full-scale IQ** | 95.9±13.7***,†† | 109.4 ±13.2 | 111.7 ±18.8 |
| **Conners Inattention** | 82.3±9.9*** | 81.8±9.4†† | 50.6±12.7 |
| **Conners Hyperactivity** | 83.4±9.4*** | 81.2±12.3†† | 51.5±13.5 |
| **Concurrent Medications:** |  |  |  |
| None | 11 | 21 | 25 |
| Methylphenidate | 5 | 3 | 0 |
| Amphetamines | 7 | 0 | 0 |
| Atomoxetine | 1 | 0 | 0 |
| Guanfacine | 1 | 0 | 0 |
| Clonidine | 1 | 0 | 0 |
| Fluoxetine | 3 | 1 | 0 |
| Risperidone | 1 | 1 | 0 |
| Aripiprazole | 2 | 0 | 0 |
| Lamotrigine | 1 | 0 | 0 |
| Valproate | 1 | 0 | 0 |
| Oxcarbazepine | 1 | 0 | 0 |
| **Prenatal Exposures:** |  |  |  |
| None | 0 | 25 | 25 |
| Ethanol | 26 | 0 | 0 |
| Tobacco | 2 | 0 | 0 |
| Cannabis | 7 | 0 | 0 |
| Stimulants | 10 | 0 | 0 |
| Opioids | 2 | 0 | 0 |
| Barbiturates | 1 | 0 | 0 |
| Cocaine | 1 | 0 | 0 |
| Unspecified | 2 | 0 | 0 |

ADHD+PAE=attention deficit hyperactivity disorder likely due to prenatal alcohol exposure; ADHD-PAE=attention deficit hyperactivity disorder without prenatal alcohol exposure, likely due to genetics; TD=typically developing control. Conners Inattention—Parent Conners Inattention T-Score; Conners Hyperactivity—Parent Conners Hyperactivity/Impulsivity T-Score; \*\*\* $p < 0.001$ vs. ADHD-PAE, †† $p < 0.01$ vs. TD.

There were no statistically significant differences in age or sex between the groups (*p* > 0.05; Table 1). Accordingly, age and sex were not included as covariates in the subsequent analyses. FSIQ differed between groups, with the ADHD+PAE group exhibiting lower IQ relative to both ADHD–PAE (*p* < 0.001) and TD (*p* < 0.01) groups. IQ was not included as a covariate since reduced IQ is a core feature of fetal alcohol spectrum disorders, and covarying for IQ could remove variance that is intrinsic to the condition.

Medication use differed across the groups, with greater medication use in the ADHD+PAE group, lower use in the ADHD–PAE group, and no medication use in the TD group. Medication use was not included as a covariate, as it reflects the clinical phenotype of the population.

### MRI Data Acquisition

All participants underwent whole-brain T1- and T2-weighted (T1w/T2w) structural MRI and resting-state functional MRI (rsfMRI) scanning on a 3 Tesla Siemens Prisma scanner using a 32-channel head coil. Data were acquired using Human Connectome Project (HCP) sequences including high-resolution motion-tolerant T1w/T2w structural MRI as in our prior work (Kilpatrick et al. 2021,2022) and a multiband echo-planar rsfMRI sequence (Smith et al. 2013). T1w, T2w, and/or rsfMRI scans were repeated when unduly contaminated by motion, as detected by automated Framewise Integrated Real-time MRI Monitoring (FIRMM; Dosenbach et al. 2017), operator inline monitoring of successive TRs, and visual observation of the participant during acquisition. For T1w MRI, scanning parameters were repetition time (TR) = 2500 ms; echo times (TEs) = 1.81, 3.6, 5.39, 7.18 ms, flip angle 8°, voxel size = 0.8 × 0.8 × 0.8 mm³. For co-registered T2w MRI, scanning parameters were TR = 3200 ms, TE = 564 ms, voxel size = 0.8 × 0.8 × 0.8 mm³. Scanning parameters for rsfMRI were TR = 800 ms, TE = 37 ms, voxel size = 2 × 2 × 2 mm³, and multiband acceleration factor = 8. Each resting-state scan lasted 13 minutes, yielding 975 time points per participant. Participants were instructed to remain still with eyes open during scanning and not to fall asleep.

### Preprocessing

T1w/T2w structural MRI and rsfMRI data were minimally preprocessed using the Human Connectome Project pipeline (Glasser et al. 2013). Briefly, the T1w/T2w pipeline includes correction of gradient distortions, cropping the neck, transformation to MNI space, brain extraction, down-sampling to FreeSurfer space, then running recon-all in FreeSurfer 6.0 to transform the cortex from its native-space volume to a native surface mesh. The rsfMRI pipeline includes the standard preprocessing steps for gradient-distortion correction, motion correction, image-distortion correction, removal of non-brain tissue, spatial normalization, and artifact removal. Any data affected by head motion (mean framewise displacement > 0.2 mm) were excluded as were frames with less than five contiguous frames of low motion data between instances of high motion (Power et al. 2014).

### Brain Parcellation

The brain was parcellated using the CAB-NP pediatric atlas (Ji et al. 2019). This parcellation draws from the Human Connectome Project-MultiModal Parcellation (HCP-MMP; Glasser et al. 2016) of 360 (180 per hemisphere) cerebral cortical regions augmented by the 66 subcortical regions of the Extended HCP parcellation (HCPex; Huang et al. 2022). The locations of the HCP-MMP and HCPex parcels are nicely viewed on the inflated brains in Rolls et al. (2022). To these CAB-NP further adds several unassigned but distinct cerebellar and brainstem sites. Each region was treated as a node in the functional connectivity analysis. CAB-NP divides these nodes into 12 functional networks (Fig. S1 Supplemental Materials). These networks include the primary visual (VIS1), secondary visual (VIS2), auditory (AUD), somatomotor (SMN), cingulo-opercular (CON), default-mode (DMN), dorsal attention (DAN), frontoparietal (FPN), posterior multimodal (PMM), ventral multimodal (VMM), orbito-affective (ORA), and language (LAN) networks.

### Functional Connectivity Analysis

For each participant, blood oxygen level–dependent (BOLD) time series were extracted from each parcellated brain region across the full duration of the resting-state scan (975 time points). Functional connectivity (fc) between each pair of regions was computed as the Pearson correlation coefficient between their respective BOLD time series. This resulted in a subject-level connectivity matrix containing pairwise connectivity values for all region pairs. Group-level connectivity values were calculated by averaging functional connectivity across participants within each group (ADHD+PAE, ADHD–PAE, and TD) for each node pair.

### Statistical Analysis

GraphVar (Kruschwitz et al. 2015) was used to conduct the network analysis. Between-group differences in functional connectivity were assessed separately for each node pair by using permutation-based testing with 10,000 permutations. Resulting p-values were corrected for multiple comparisons using the false discovery rate (FDR; Benjamini & Yekutieli 2001) with a significance threshold of *q* < 0.05.

Pairwise group comparisons were performed first for ADHD+PAE versus ADHD–PAE across the brain to identify networks containing edges with significant differences between the two ADHD etiologies. Then, for each network containing such edges, ADHD-PAE and ADHD+PAE were separately compared to TD. to aid in interpretation of findings. Both directions of effect (greater than and less than) were evaluated for each comparison.

### Visualization

First, for each functional network showing edges with significant ADHD+PAE versus ADHD–PAE fc differences, the corresponding partial matrix of the whole-brain fc matrix was displayed. This was accompanied by a lateral-view transparent surface rendering (glass brain) within which the edges with significant difference were depicted. Then, for each network, significant differences between groups in functional connectivity were visualized using circular connectograms. Each node represents a brain region from the CAB-NP atlas, and edges represent node pairs showing statistically significant group differences in functional connectivity. Separate connectograms were generated for each pairwise comparison and direction of effect (e.g., ADHD+PAE > ADHD–PAE, ADHD+PAE < ADHD–PAE, etc.). The edge colors reflect anatomical groupings of regions, including frontal cortex, parietal/posterior cingulate/temporal cortex, cerebellum, and subcortical regions, as defined in the labeling scheme (Figs. 1-2).

**Fig. 1.**
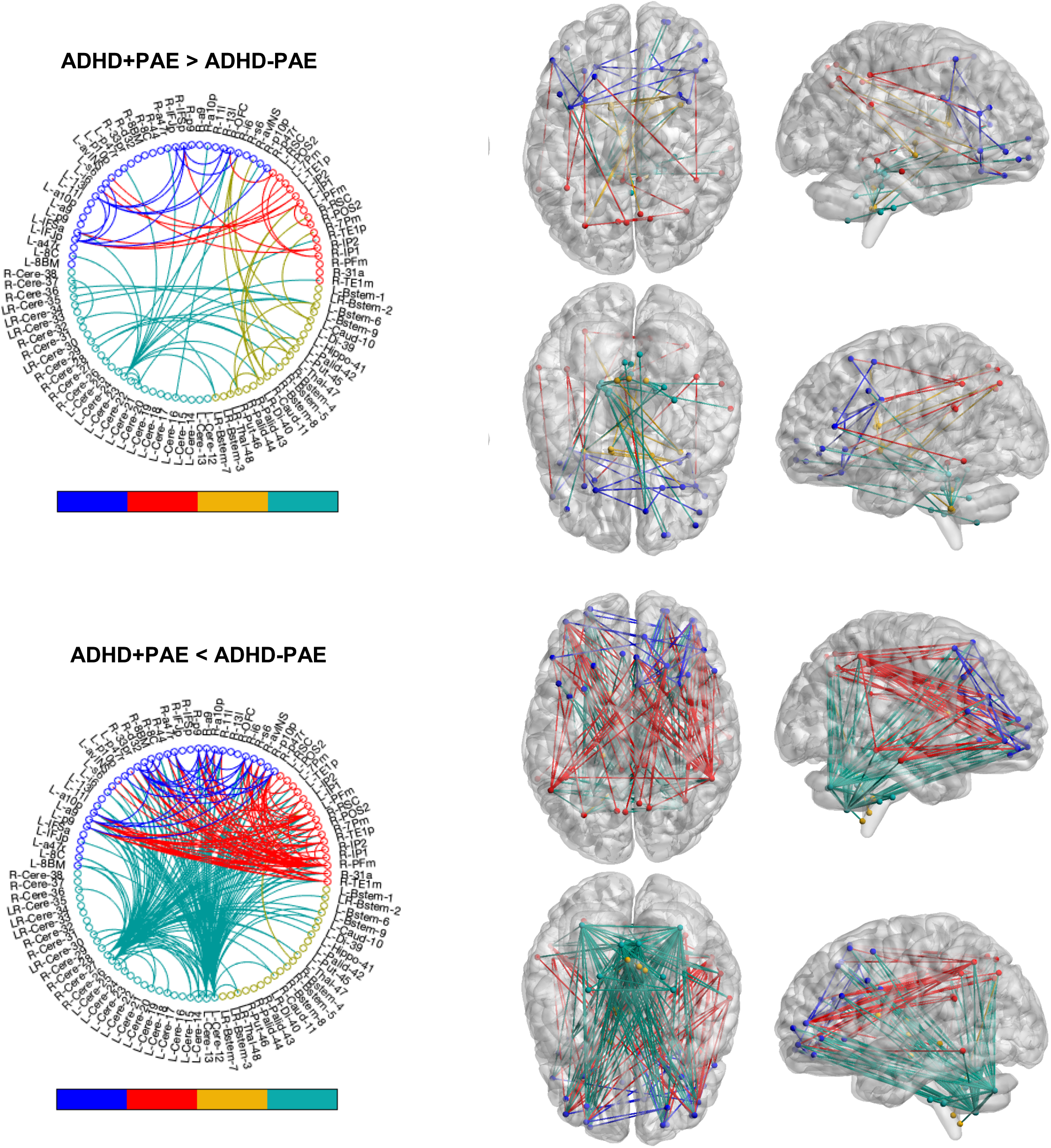
Differences in resting-state functional connectivity (fc) within the frontoparietal network (FPN) between two etiologies of pediatric ADHD. The two etiologies are: ADHD likely due to confirmed prenatal alcohol exposure (ADHD+PAE) and probable familial ADHD with no evidence of prenatal alcohol exposure (ADHD–PAE). Differences are shown on circular connectograms of the 76 neuroanatomic nodes of the FPN (*left*) and on brain surface renderings (“glass brains”, *right*). Glass brains are seen in dorsal (*center right upper*), basal (*center right lower*), right lateral (*center right upper*), and left lateral (*center right lower*) view. For both connectograms and glass brains, each link displayed represents a connection between an anatomic node pair (edge) within the FPN with a significant between-group difference (10,000 permutations, FDR-corrected, *q* < 0.05) in mean fc. (*Upper*) links with higher fc in ADHD+PAE than in ADHD-PAE; (*lower*) links with lower fc in ADHD+PAE than in ADHD-PAE. Although a number of edges show higher fc in ADHD+PAE, a majority have lower fc than in ADHD-PAE in this network, including many connections with the cerebellum. Significant between-group differences were not found in other networks. Figures prepared with GraphVar (Kruschwitz et al. 2015). Nodes represent CAB-NP (Ji et al. 2019) regions color-coded by anatomical group (frontal-- *blue*, parietal-cingulate-temporal-- *red*, subcortical—*orange-green* or *orange*, cerebellar-- *teal*). (For display purposes in the glass brains, all the distinct cerebellar nodes in the connectograms are collapsed into a single point for each cerebellum and the left cerebellum is rendered invisible.)

**Fig. 2.**
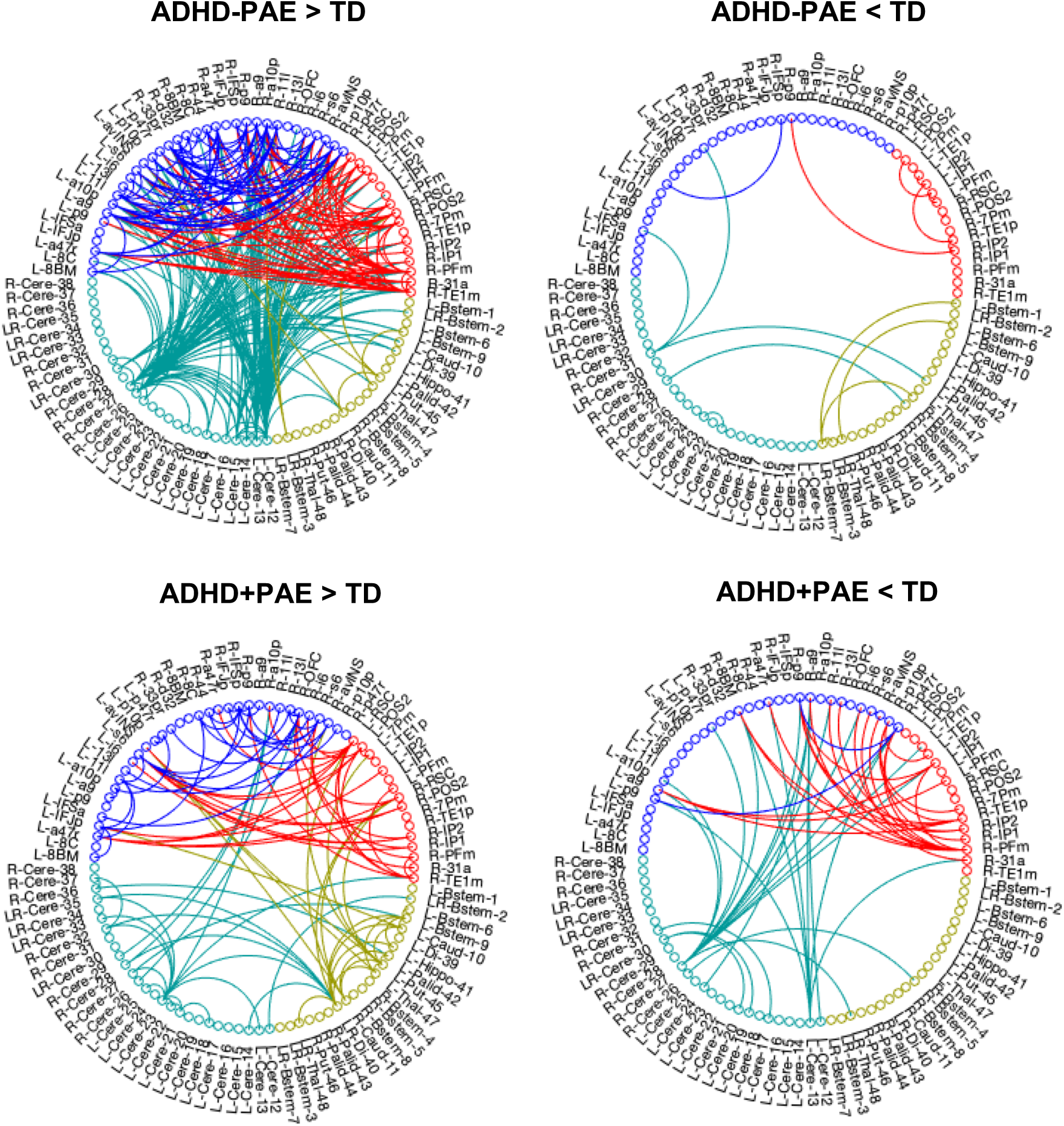
Circular connectograms comparing resting-state functional connectivity (fc) within the frontoparietal network (FPN) between two etiologies of pediatric ADHD and typically developing (TD) children. The two etiologies are ADHD likely due to confirmed prenatal alcohol exposure (ADHD+PAE) and probable familial ADHD with no evidence of prenatal alcohol exposure (ADHD–PAE). Differences are shown on circular connectograms of the 76 neuroanatomic nodes of the FPN Each link displayed represents a connection between an anatomic node pair (edge) within the FPN with a significant between-group difference (10,000 permutations, FDR-corrected, *q* < 0.05) in mean fc. (*Upper*) links with higher (*left*) or lower (*right*) fc in ADHD-PAE than in TD; (*lower*) links with higher (*left*) or lower (*right*) fc in ADHD+PAE than in TD. Nodes represent CAB-NP (Ji et al. 2019) regions and are color-coded by anatomical group frontal-- *blue*, parietal-cingulate-temporal-- *red*, subcortical—*orange-green*, cerebellar-- *teal*). For ADHD-PAE, there are numerous FPN edges with hyperconnectivity and very few with hypoconnectivity with respect to TD; for ADHD+PAE, the picture is more mixed but still with a majority of hyperconnected edges with respect to TD.

## 3. Results

### Connectome-wide analysis of ADHD+PAE versus ADHD–PAE

Permutation-based connectome-wide analyses (FDR-corrected *q* < 0.05) identified widespread group differences in resting-state functional connectivity (fc) across the CAB-NP pediatric parcellation that were, however, restricted to edges connecting nodes within the frontoparietal network (FPN). These are shown in the circular connectograms and brain renderings for each contrast involving ADHD+PAE and ADHD-PAE (Fig. 1). After correction for multiple comparisons, edges with significant differences between the two ADHD etiologies were not found within other networks or between networks. Fig. S1 (Supplementary Materials) shows the FPN for the CAB-NP Atlas and Table S1 (Supplementary Materials) lists its 76 neuroanatomic regions (nodes), as labelled in the connectograms (Figs. 1-2).

### ADHD+PAE versus ADHD-PAE breakdown of significant node-pair connections

ADHD+PAE exhibited lower fc than ADHD–PAE across 234 unique edges (Fig. 1, Table S2b Supplementary Materials), with the largest share involving cerebellar–cerebral cortical connections (110/234), followed by cerebral cortical–cerebral cortical connections (85/234) and cerebellar–cerebellar connections (23/234), with additional cerebellar–subcortical/brainstem connections (15/234). The strongest hubs in this contrast were cerebellar regions, including L-Cere-12 (degree = 45), R-Cere-26 (degree = 37), R-Cere-27 (degree = 35), and L-Cere-13 (degree = 31), alongside an association cortical hub R-PFm (degree = 24).

A smaller set of edges showed higher fc in ADHD+PAE than ADHD–PAE (67 unique edges) (Fig. 1, Table S2a Supplementary Materials). These effects included cerebral cortical–cerebral cortical connections (26/67) and multiple cross-structure connections involving cerebellum–subcortical/brainstem (12/67), cerebellum–neocortex (10/67), and neocortex–subcortical/brainstem (8/67). High-degree nodes in this set included L-Cere-23 (degree = 13), L-IFJp (degree = 7), R-IFSp (degree = 6), and L-Bstem-6 (degree = 6).

### ADHD–PAE versus typically developing controls (TD) breakdown of significant node-pair connections

Children with ADHD–PAE showed higher fc than TD across 255 unique edges (Fig. 2, Table S3a Supplementary Materials). These effects were distributed across cerebral cortical–cerebral cortical connections (117/255) and cerebellar–cerebral cortical connections (97/255), with additional cerebellar–cerebellar (22/255) and cerebellar–subcortical/ brainstem (12/255) differences. The most highly connected hubs in this contrast included R-Cere-26 (degree = 42) and L-Cere-12 (degree = 38), along with association cortical nodes including R-33pr (degree = 20), R-a10p (degree = 19), R-TE1m (degree = 18), and R-PFm (degree = 17).

A smaller set of edges showed lower fc in ADHD–PAE than TD (16 unique edges) (Fig. 2, Table S3b Supplementary Materials). These differences spanned cerebral cortical–cerebral cortical (7/16), subcortical/brainstem (4/16), and cerebellar-related connections (5/16 combined cerebellar–cortical, cerebellar–subcortical/brainstem, and cerebellar–cerebellar).

### ADHD+PAE vs typically developing controls (TD)

Relative to TD, children with ADHD+PAE showed both increased and decreased connectivity (Fig. 2, Table S4 Supplementary Materials). ADHD+PAE demonstrated higher fc than TD across 105 unique edges, primarily involving cerebral cortical–cerebral cortical connections (56/105), with additional differences in subcortical/brainstem–subcortical/brainstem (14/105), cerebral cortical–subcortical/brainstem (10/105), and cerebellar-related connections (25/105 combined cerebellar–cerebellar, cerebellar–cerebral cortical, and cerebellar–subcortical/brainstem). Prominent nodes in this hyperconnectivity pattern included LR-Di-40 (degree = 11) and L-Cere-23 (degree = 11), along with L-s6 (degree = 9), R-IFSp (degree = 8), L-Put-45 (degree = 8), and L-RSC (degree = 8).

In contrast, ADHD+PAE showed lower fc than TD across 47 unique edges (Fig. 2, Table S4 Supplementary Materials). These reductions were concentrated in cerebral cortical–cerebral cortical (23/47) and cerebellar–cerebral cortical (19/47) connections, with smaller numbers of cerebellar–cerebellar (3/47) and cerebellar–subcortical/brainstem (2/47) differences. Nodes with the highest degree in this hypoconnectivity set included R-Cere-27 (degree = 12) and R-PFm (degree = 8), followed by L-Cere-13 (degree = 6), R-p47r (degree = 6), R-a9 (degree = 5), and R-p9 (degree = 5).

### Overlap across contrasts

Across contrasts, the ADHD+PAE hypoconnectivity pattern relative to TD substantially overlapped with the ADHD+PAE hypoconnectivity pattern relative to ADHD–PAE: 36 of 47 ADHD+PAE < TD edges were also present in ADHD+PAE<ADHD–PAE. In addition, 121 edges were shared between ADHD–PAE > TD and ADHD+PAE < ADHD–PAE. Only one edge overlapped between the two TD hypoconnectivity contrasts (ADHD+PAE < TD and ADHD–PAE < TD), and six edges showed opposite-direction differences between ADHD+PAE and ADHD–PAE when each was compared to TD.

## 4. Discussion

The major findings of this study were: (1) children with ADHD+PAE showed widespread lower functional connectivity (fc) than children with ADHD–PAE in the frontoparietal network (FPN) with reductions concentrated primarily in cerebellar–cerebral cortical and cerebral cortical–cerebral cortical connections, (2) children with ADHD–PAE showed widespread hyperconnectivity of FPN regions relative to typically developing (TD) controls, and (3) children with ADHD+PAE showed a mixed pattern relative to TD (many hyperconnected region pairs but also a moderate number of hypoconnected pairs). Taken together, these results suggest that ADHD, in the context of prenatal alcohol exposure is neurally distinct from ADHD without PAE, and that the presence of PAE is associated with relatively reduced large-scale connectivity, especially across cerebellar–cerebral cortical systems.

The first major finding was that children with ADHD+PAE showed substantially lower fc than children with ADHD–PAE. This was the strongest contrast in the study, with 234 edges showing reduced connectivity in ADHD+PAE relative to ADHD–PAE and only 67 edges showing increased connectivity. The reduced edges were concentrated especially in cerebellar–cerebral cortical and cerebral cortical–cerebral cortical connections and the highest-degree hubs in this pattern were primarily cerebellar, including L-Cere-12, R-Cere-26, R-Cere-27, and L-Cere-13, along with the association cortical node R-PFm. This finding is important since it indicates that ADHD+PAE is not just a more severe version of ADHD–PAE. Instead, the direct comparison showed a different profile, with ADHD+PAE exhibiting reduced connectivity across many of the same systems in which ADHD–PAE showed increased connectivity relative to TD. This supports the view that ADHD+PAE and ADHD–PAE represent distinct neural phenotypes (Ding et al. 2021). The prominence of cerebellar hubs in both the ADHD–PAE > TD and ADHD+PAE < ADHD–PAE contrasts also fits with the growing view that cerebellar dysfunction is relevant to ADHD pathophysiology, including not only motor regulation but also cognitive and attentional functions (Isaac et al. 2025).

The second major finding was that children with ADHD–PAE showed widespread hyperconnectivity relative to TD, with 255 edges showing higher resting-state functional connectivity (fc) and only 16 showing lower fc. This pattern was broadly distributed and involved cerebral cortical–cerebral cortical, cerebellar–cerebral cortical, cerebellar–cerebellar, and cerebellar–subcortical/brainstem connections. The strongest hubs in this contrast included cerebellar nodes such as R-Cere-26 and L-Cere-12, along with association cortical nodes including R-33pr, R-a10p, R-TE1m, and R-PFm. This finding indicates that ADHD–PAE was characterized primarily by increased fc relative to typical development. This interpretation is broadly consistent with prior ADHD connectivity work showing that ADHD is associated with complex alterations in distributed functional networks rather than focal abnormalities alone (Zhao et al. 2022), and with EEG and resting-state work specifically in frontal, central and medial temporal regions in children with ADHD (Chen et. al. 2021, Cortese et. al. 2012).

The third major finding was that children with ADHD+PAE showed a mixed connectivity pattern relative to TD, with 105 edges showing higher fc and 47 showing lower fc. Although more edges showed higher fc than lower fc in ADHD+PAE relative to TD, the pattern was not as uniformly hyperconnected as in ADHD–PAE. Increased fc in ADHD+PAE involved cerebral cortical–cerebral cortical, subcortical/brainstem, cerebral cortical–subcortical/brainstem, and cerebellar-related connections, with prominent nodes including LR-Di-40, L-Cere-23, L-s6, R-IFSp, L-Put-45, and L-RSC. In contrast, reduced fc in ADHD+PAE relative to TD was concentrated mainly in cerebral cortical–cerebral cortical and cerebellar–cortical connections, with hubs including R-Cere-27 and R-PFm. Thus, unlike ADHD–PAE, ADHD+PAE was not characterized by a predominantly hyperconnected profile. Instead, it showed both increases and decreases in fc, suggesting a less uniform and more regionally variable pattern of network alteration. This mixed pattern is compatible with prior work showing that ADHD-related connectivity differences are heterogeneous and can involve more complex distributed covariance structures than simple one-directional increases or decreases (Castellanos & Proal 2012).

An additional important finding was the degree of overlap across contrasts. Of the 47 edges showing lower fc in ADHD+PAE relative to TD, 36 were also present in the ADHD+PAE < ADHD–PAE contrast. In addition, 121 edges were shared between ADHD–PAE > TD and ADHD+PAE < ADHD–PAE. This overlap strengthens the interpretation that hypoconnectivity is a defining feature of ADHD+PAE relative to both of the comparison groups. It also suggests that many of the connections that distinguish ADHD–PAE from typical development in the direction of increased fc are the same as the connections that separate ADHD+PAE from ADHD–PAE in the reduced fc direction. In contrast, there was minimal overlap between the two TD hypoconnectivity contrasts as they had only one shared edge. Together, these overlap analyses further support the conclusion that ADHD+PAE is not simply an intermediate between ADHD–PAE and TD, but instead shows a partially distinct connectivity signature.

These findings are consistent with the hypothesis that ADHD–PAE would show a predominantly hyperconnected profile relative to TD, whereas ADHD+PAE would be more underconnected. The hypothesis was only partially supported because ADHD–PAE did show the expected predominantly hyperconnected pattern but ADHD+PAE did not show uniformly reduced fc relative to TD, but instead a mixed pattern of increases and decreases. Still, the direct comparison with ADHD–PAE, together with the overlap analyses, showed that ADHD+PAE was characterized by substantial reductions in connectivity, especially across cerebellar–cortical and cortical–cortical edges. Thus, the broader prediction that ADHD+PAE would be neurally distinct from ADHD–PAE and relatively less connected across major systems was supported.

### 4.1. Limitations

This present study has limitations. The sample size was modest though similar to previous ADHD and PAE studies involving rsfMRI (Aoki et. al. 2018, Ghazi Sherbaf et. al. 2019). Racial diversity is also a shortcoming, as there was a majority of white participants, no Black TD, and only one Hispanic ADHD-PAE participant. Mean FSIQ was lower in the ADHD+PAE sample than in the other two samples, although all children did have FSIQ in the normal range (> 70). As mentioned, we did not covary for FSIQ as low IQ is highly associated with PAE. As children with PAE typically undergo psychopharmacologic treatment, current use of psychiatric medications was more common in the ADHD+PAE sample than in the other samples. Also, beyond ethanol, many participants in the ADHD+PAE sample were exposed to other drugs-of-abuse *in utero* (Table 1). This is, unfortunately, very common in the PAE population; to exclude such participants, or to exclude medicated participants, would represent substantial recruitment challenge and would be clinically unrealistic.

### 4.2. Conclusions

Notwithstanding these limitations, this study reveals that there are notable differences in cerebral and cerebellar resting-state functional connectivity between ADHD+PAE, ADHD-PAE, and typically developing controls. A clinically relevant future step of this research would be to determine whether the BOLD fc profile can be used to perform differential diagnosis of ADHD+PAE vs. ADHD-PAE on a single-subject level.

## Data Availability

All data produced in the present study are available upon reasonable request to the authors

## Acknowledgements

We wish to thank the patients and families who participated in this research.

## SUPPLEMENTARY MATERIALS

**Fig. S1.**
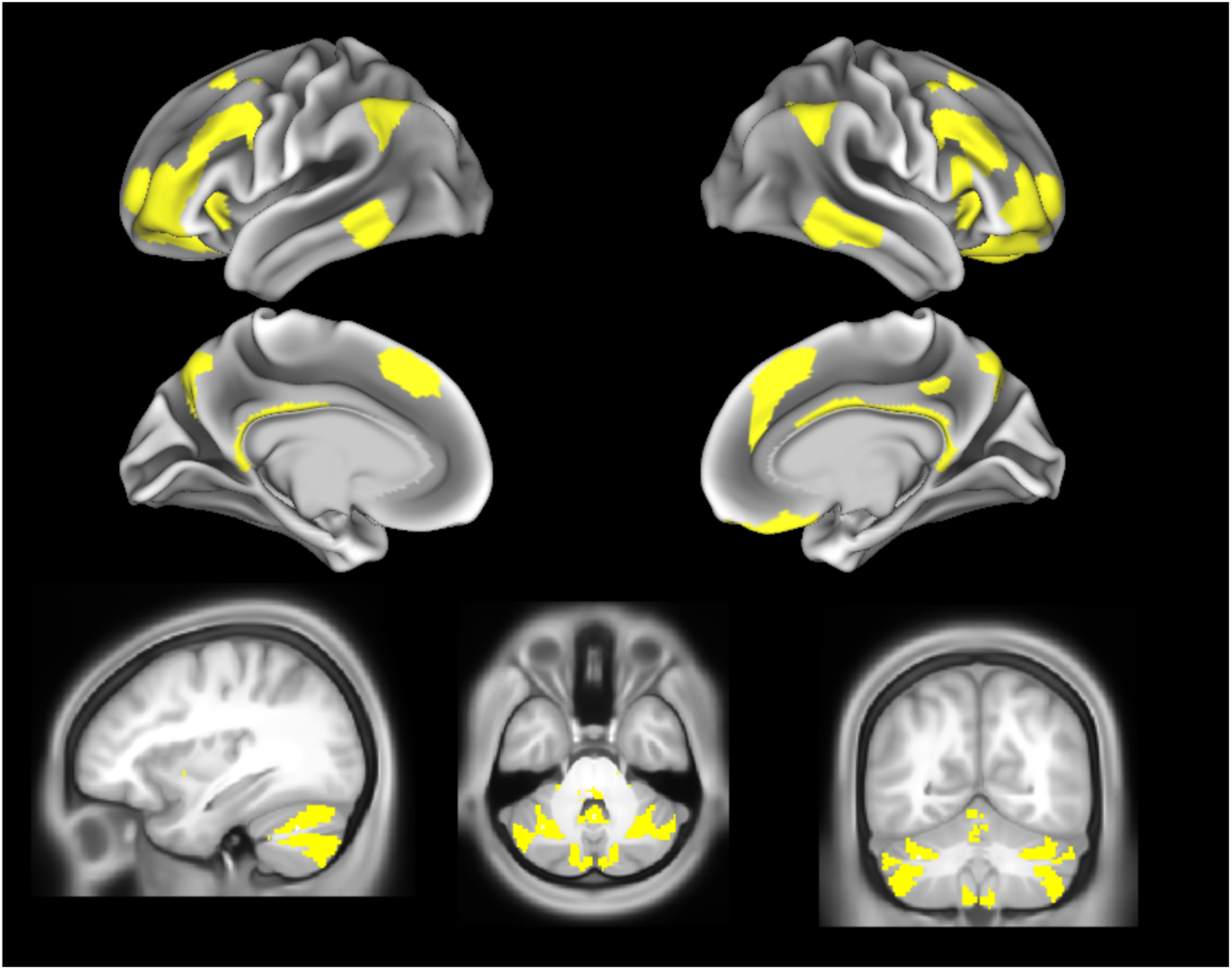
The frontoparietal network (FPN) in the Cole-Anticevic Brainwide Network Parcellation (CAB-NP) pediatric brain atlas (Ji et al. 2019). (*Upper*) cortical surface renderings of lateral (*uppermost*) and mesial (*upper central*) views of the left (*left*) and right (*right*) cerebral hemispheres. (*Lower*) parasagittal (*left*), transverse (*center*), and coronal (*right*) T1-weighted MRI sections to expose the cerebella. Regions assigned to the FPN appear in *yellow*. Note numerous cerebellar nodes within the CAB-NP FPN.

**Table S1a.** Frontoparietal network (FPN) nodes in the Cole-Anticevic Brainwide Network Parcellation (CAB-NP) Atlas (Ji et al. 2019). Nodes in original Human Connectome Project-MultiModal Parcellation (HCP-MMP; Glasser et al. 2016)

| Node Name | Area Description | Other Names | Gross Location | Glasser Parcel# |
| --- | --- | --- | --- | --- |
| 8BM | Area 8BM | 8B | mesial premotor cortex | 63 |
| 8C | Area 8C | 8Av | frontal eye fields | 73 |
| a47r | Area anterior 47r | 47r | orbitofrontal cortex | 77 |
| IFJp | Area IFJp |  | posterior inferior frontal junction | 80 |
| IFSa | Area IFSa |  | anterior inferior frontal sulcus | 82 |
| p9(-46v) | Area posterior 9-46v | 9-46v | middle frontal cortex | 83 |
| a9(-46v) | Area anterior 9-46v | 9-46v | middle frontal cortex | 85 |
| a10p | Area anterior 10p | 10p, 10, Fp1 | frontal polar cortex | 89 |
| 11l | Area 11l | Fo3 | orbitofrontal cortex | 91 |
| 13l | Area 13l | Fo3 | orbitofrontal cortex | 92 |
| i6(-8) | Inferior 6-8 Transitional Area | FC(B) | premotor cortex | 97 |
| s6 | Superior 6-8 Transitional Area | FC(B) | supplementary motor cortex | 98 |
| avI(NS) | Anterior Ventral Insular Area | lai | anterior insula | 111 |
| p10p | Area posterior 10p | 10p, 10, Fp1 | frontal polar cortex | 170 |
| p47r | Area posterior 47r | 47r | orbitofrontal cortex | 171 |
| 33pr | Area 33 prime | 33', 16 | anterior middle cingulate cortex | 58 |
| d32 | Area dorsal 32 | 32 | anterior middle cingulate cortex | 62 |
| 44 | Area 44 | 44d, 44v | inferior frontal cortex pars opercularis | 74 |
| IFSp | Area IFSp |  | posterior inferior frontal junction | 81 |
| OFC | Orbital Frontal Complex | 11m, 13b, 13m, 14r, Fo1 | orbitofrontal cortex | 93 |
| RSC | RetroSplenial Complex | 29, 30 | retrosplenial cortex | 14 |
| POS2 | Parieto-Occipital Sulcus Area 2 |  | retrosplenial cortex | 15 |
| 7Pm | Medial Area 7P | 7P | precuneus | 29 |
| TE1p | Area TE1 posterior |  | superior temporal cortex | 133 |
| IP2 | Area IntraParietal 2 |  | intraparietal sulcal cortex | 144 |
| IP1 | Area IntraParietal 1 |  | intraparietal sulcal cortex | 145 |
| PFm | Area PFm Complex | 40, 89 | supramarginal cortex | 149 |
| 31a | Area 31a | 31, 31d, 31v | dorsal posterior cingulate cortex | 162 |
| TE1m | Area TE1 Middle |  | superior temporal cortex | 177 |

**Table S1b.**
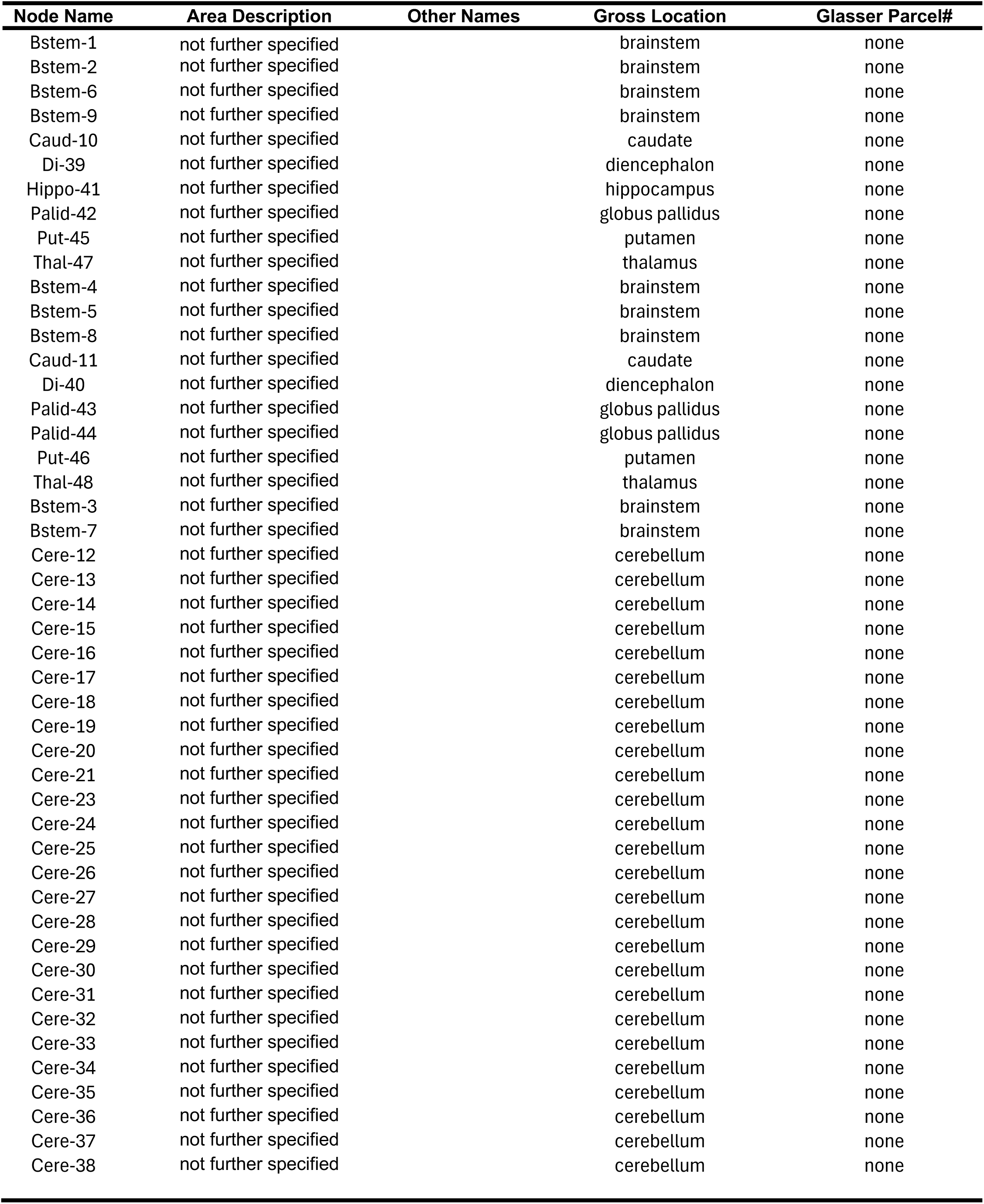
Subcortical nodes added in the Extended HCP parcellation (HCPex; Huang et al. 2022) and distinct but unspecified brainstem and cerebellar nodes added by CAB-NP (Ji et al. 2019).

**Table S2a.** CAB-NP node pairs with higher resting-state functional connectivity in ADHD+PAE than in ADHD-PAE.

| Node 1 | Node 2 | Node 1 | Node 2 |
| --- | --- | --- | --- |
| L-8C | L-Cere-23 | L-Bstem-6 | L-Cere-23 |
| L-IFJp | L-IFSa | L-Bstem-6 | LR-Cere-34 |
| L-IFJp | L-13l | L-Bstem-6 | LR-Cere-35 |
| L-IFJp | L-s6 | L-Palid-42 | R-Caud-11 |
| L-IFJp | R-IFSp | L-Palid-42 | R-Cere-36 |
| L-IFJp | R-p47r | L-Put-45 | L-Thal-47 |
| L-IFJp | L-7Pm | L-Put-45 | R-Cere-28 |
| L-IFJp | L-IP2 | L-Thal-47 | R-Put-46 |
| L-IFSa | L-i6 | R-Bstem-5 | R-Cere-28 |
| L-IFSa | L-TE1p | R-Bstem-5 | R-Cere-29 |
| L-IFSa | L-Cere-23 | LR-Di-40 | LR-Thal-48 |
| L-13l | R-IFJp | LR-Di-40 | L-Cere-23 |
| L-13l | R-IFSp | R-Put-46 | L-Cere-23 |
| L-13l | R-13l | LR-Bstem-7 | L-Cere-23 |
| L-13l | R-OFC | LR-Bstem-7 | R-Cere-32 |
| L-i6 | L-IP2 | L-Cere-21 | L-Cere-22 |
| L-s6 | R-IP2 | L-Cere-21 | L-Cere-23 |
| L-avINS | L-IP1 | L-Cere-21 | L-Cere-24 |
| L-p47r | L-Cere-23 | R-Cere-32 | R-Cere-37 |
| R-a47r | L-Cere-23 | R-Cere-36 | R-Cere-37 |
| R-IFJp | R-IFSp |  |  |
| R-IFJp | R-i6 |  |  |
| R-IFSp | R-p9 |  |  |
| R-IFSp | R-13l |  |  |
| R-IFSp | R-IP2 |  |  |
| R-a10p | L-Cere-23 |  |  |
| R-13l | R-OFC |  |  |
| R-OFC | L-RSC |  |  |
| R-OFC | L-Cere-16 |  |  |
| R-OFC | R-Cere-26 |  |  |
| R-i6 | L-Caud-10 |  |  |
| R-i6 | L-Put-45 |  |  |
| R-i6 | R-Caud-11 |  |  |
| R-avINS | L-Thal-47 |  |  |
| R-p10p | L-Cere-23 |  |  |
| L-RSC | R-IP1 |  |  |
| L-POS2 | R-IP1 |  |  |
| L-IP1 | R-POS2 |  |  |
| L-IP1 | R-Caud-11 |  |  |
| L-IP1 | LR-Di-40 |  |  |
| R-RSC | LR-Di-40 |  |  |
| R-7Pm | L-Caud-10 |  |  |
| R-TE1p | L-Cere-23 |  |  |
| R-TE1m | L-Cere-23 |  |  |
| L-Bstem-6 | L-Bstem-9 |  |  |
| L-Bstem-6 | R-Put-46 |  |  |
| L-Bstem-6 | L-Cere-21 |  |  |

**Table S2b.**
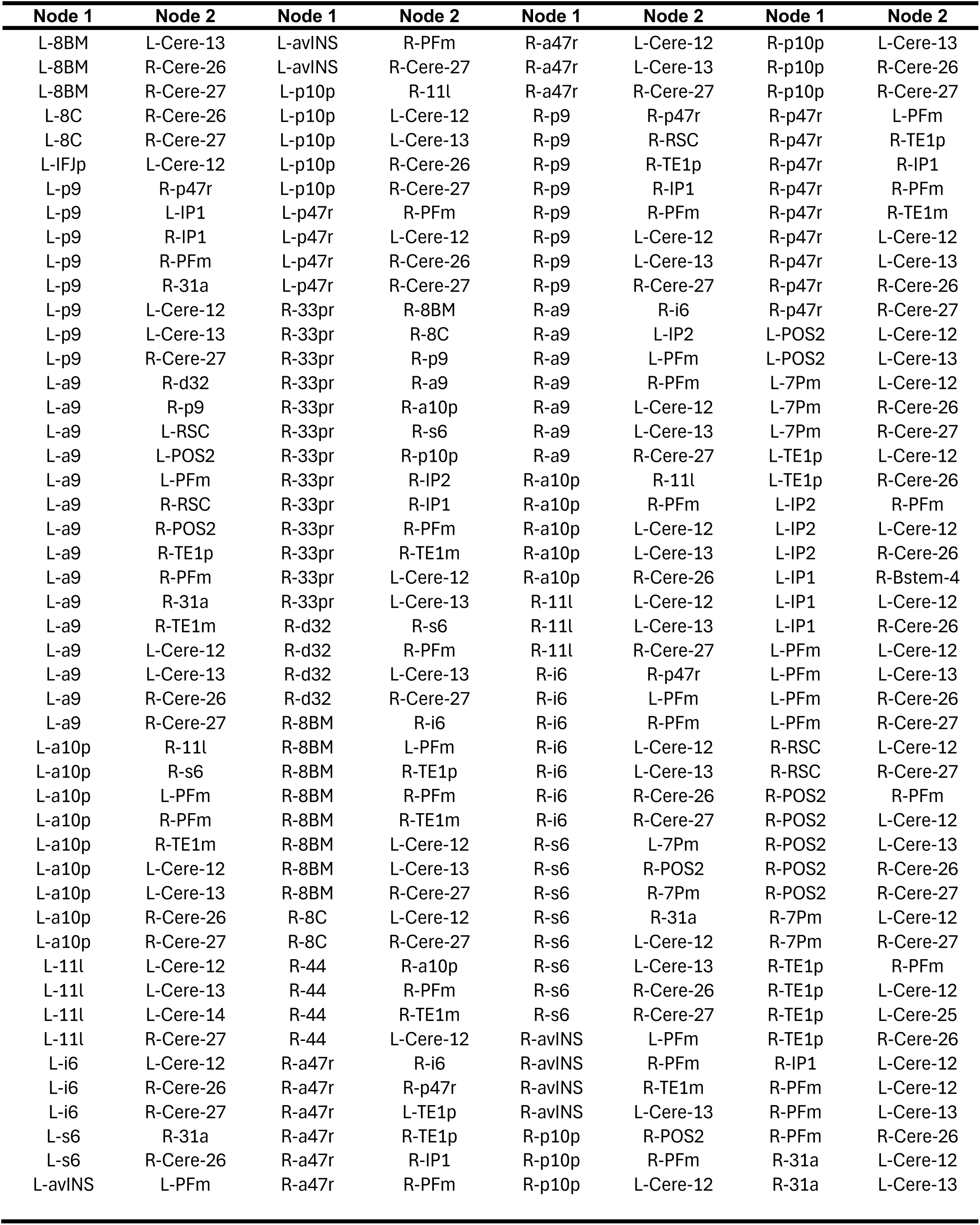
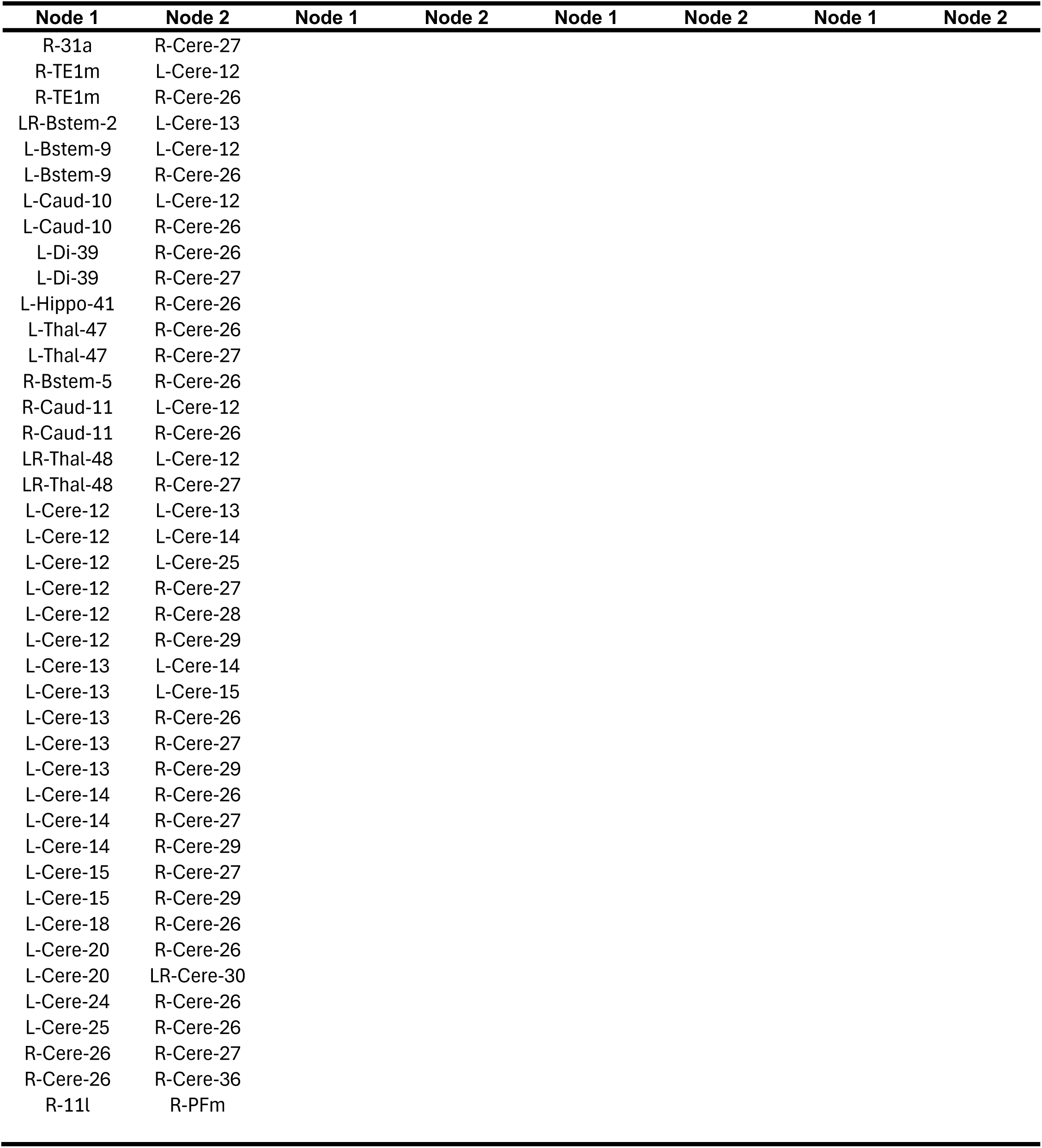
CAB-NP node pairs with lower resting-state functional connectivity in ADHD+PAE than in ADHD-PAE.

**Table S3a.**
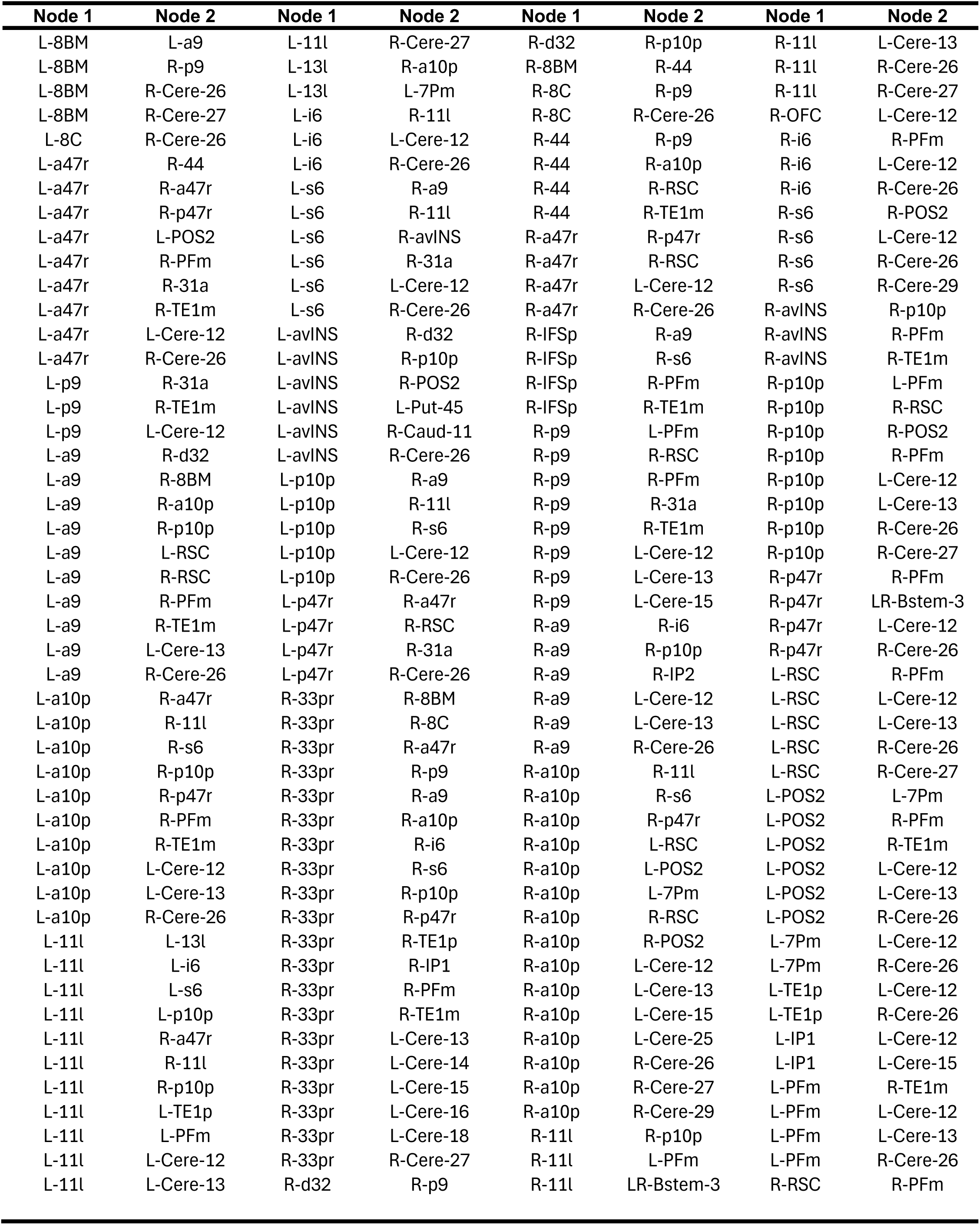
CAB-NP node pairs with higher resting-state functional connectivity in ADHD-PAE than in TD.

**Table S3b.**
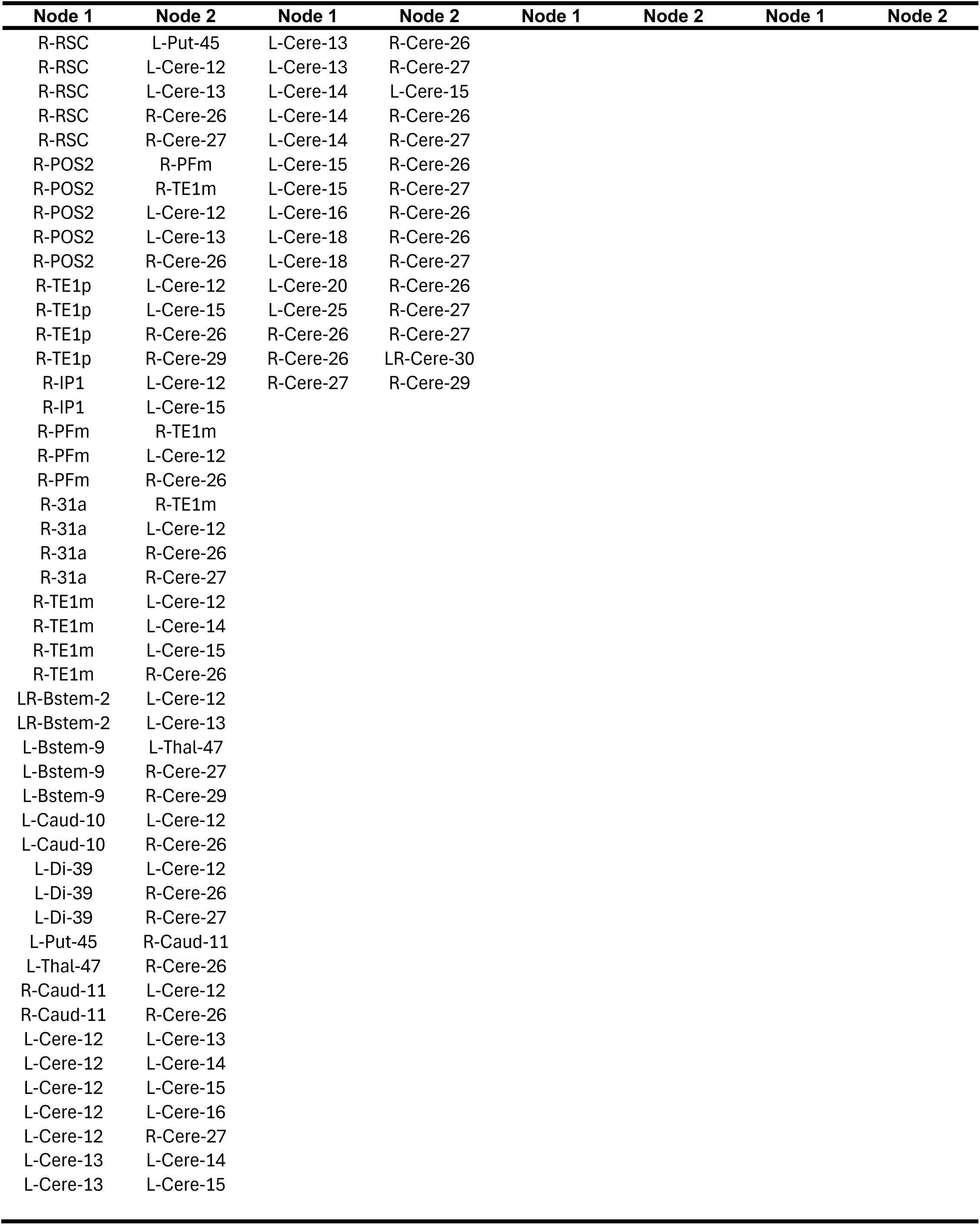
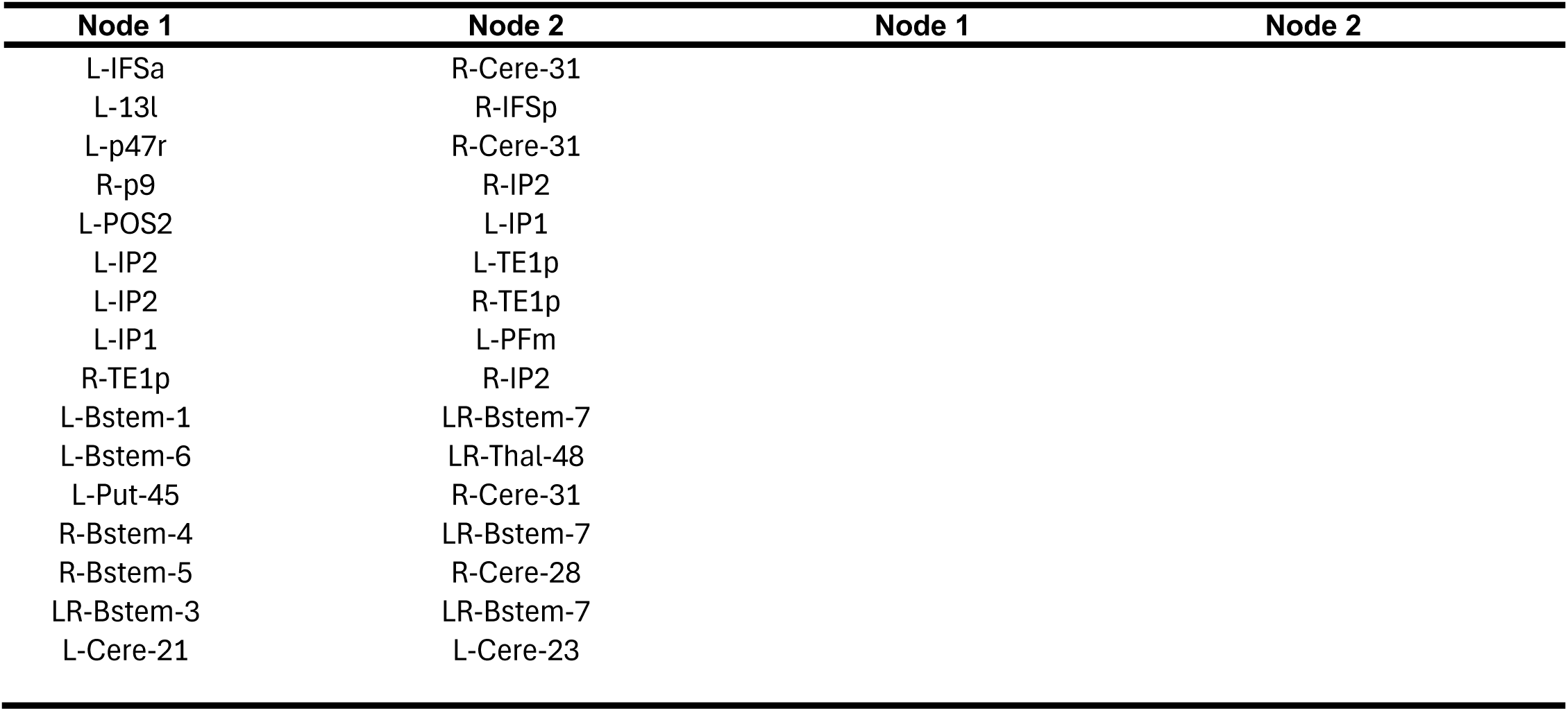
CAB-NP node pairs with lower resting-state functional connectivity in ADHD-PAE than in TD.

**Table S4a.**
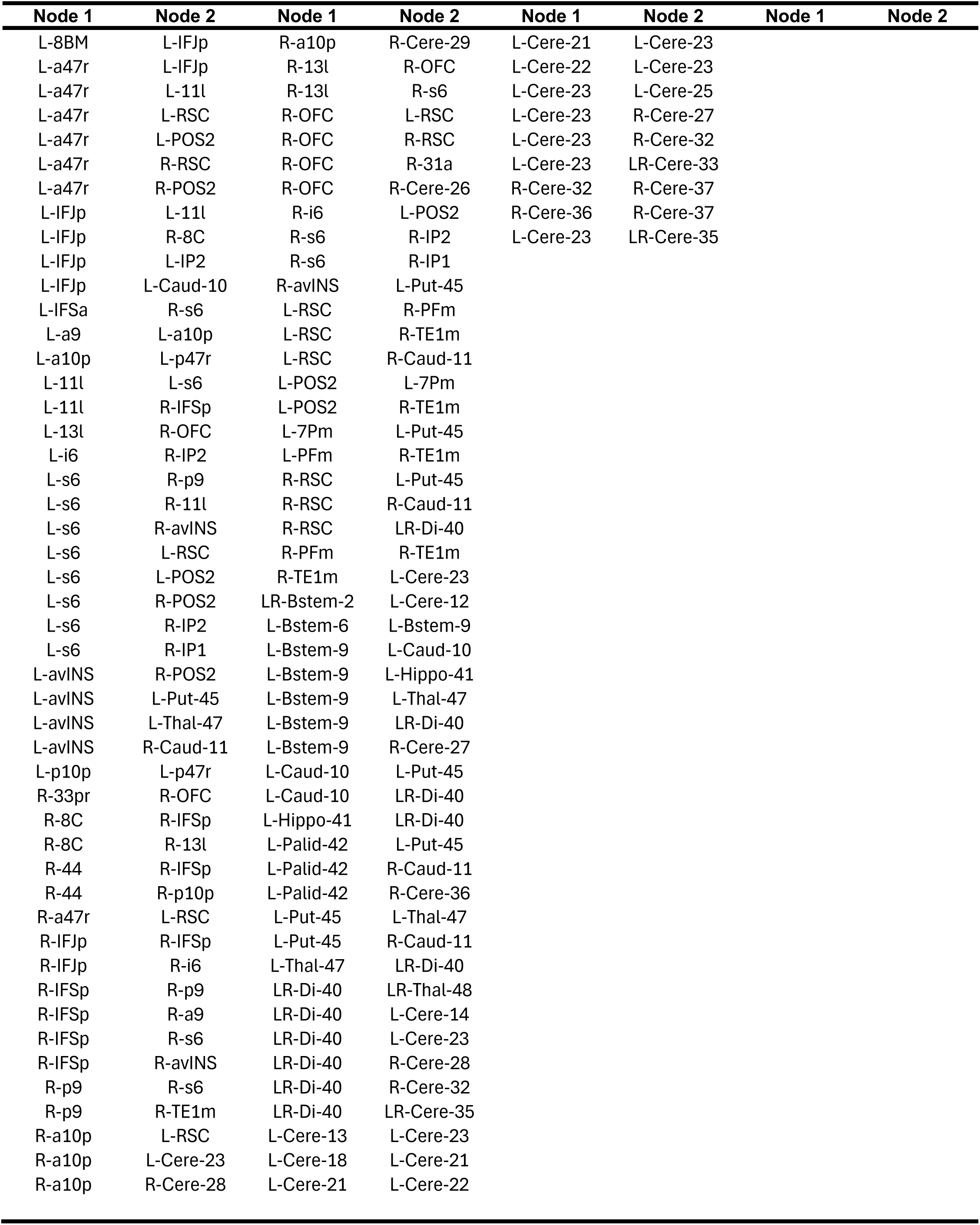
CAB-NP node pairs with higher resting-state functional connectivity in ADHD+PAE than in TD.

**Table S4b.** CAB-NP node pairs with lower resting-state functional connectivity in ADHD+PAE than in TD.

| **Node 1** | **Node 2** | **Node 1** | **Node 2** |
| --- | --- | --- | --- |
| L-p9 | R-p47r |  |  |
| L-p9 | L-Cere-13 |  |  |
| L-a9 | L-IP2 |  |  |
| L-a9 | L-PFm |  |  |
| L-a9 | R-TE1p |  |  |
| L-a9 | R-Cere-27 |  |  |
| L-a10p | R-Cere-27 |  |  |
| L-p47r | R-Cere-27 |  |  |
| R-8BM | R-TE1p |  |  |
| R-8BM | R-PFm |  |  |
| R-8BM | L-Cere-13 |  |  |
| R-8BM | R-Cere-27 |  |  |
| R-a47r | R-PFm |  |  |
| R-a47r | L-Cere-13 |  |  |
| R-p9 | R-p47r |  |  |
| R-p9 | L-IP2 |  |  |
| R-p9 | R-TE1p |  |  |
| R-p9 | R-IP2 |  |  |
| R-p9 | R-Cere-27 |  |  |
| R-a9 | R-PFm |  |  |
| R-a9 | L-Cere-12 |  |  |
| R-a9 | L-Cere-13 |  |  |
| R-a9 | L-Cere-24 |  |  |
| R-a9 | R-Cere-27 |  |  |
| R-11l | R-PFm |  |  |
| R-11l | R-Cere-27 |  |  |
| R-13l | R-IP2 |  |  |
| R-i6 | R-PFm |  |  |
| R-i6 | R-Cere-27 |  |  |
| R-s6 | R-31a |  |  |
| R-avINS | R-PFm |  |  |
| R-p10p | L-TE1p |  |  |
| R-p10p | R-7Pm |  |  |
| R-p10p | R-TE1p |  |  |
| R-p47r | R-IP1 |  |  |
| R-p47r | R-PFm |  |  |
| R-p47r | L-Cere-13 |  |  |
| R-p47r | R-Cere-27 |  |  |
| L-7Pm | R-Cere-27 |  |  |
| L-IP2 | R-PFm |  |  |
| L-IP2 | R-Cere-27 |  |  |
| R-31a | L-Cere-13 |  |  |
| R-Bstem-5 | R-Cere-26 |  |  |
| LR-Thal-48 | R-Cere-27 |  |  |
| L-Cere-17 | R-Cere-31 |  |  |
| L-Cere-19 | LR-Cere-30 |  |  |
| L-Cere-20 | LR-Cere-30 |  |  |

